# ‘Citizens’ Attitudes Under Covid19’: a cross-country panel survey of public opinion in 11 democracies

**DOI:** 10.1101/2021.09.08.21263310

**Authors:** Sylvain Brouard, Martial Foucault, Elie Michel, Michael Becher, Pavlos Vasilopoulos, Pierre-Henri Bono, Nicolas Sormani

## Abstract

This article introduces data collected in the Citizens’ Attitudes Under Covid-19 Project (CAUCP), which surveyed public opinion throughout the Covid-19 pandemic in 11 countries between March to December 2020. In this paper, we present a unique cross-country panel survey of citizens’ attitudes and behaviors during a worldwide unprecedented health, governance, and economic crisis. This dataset allows to examine the behavioral and attitudinal consequences of crisis across time and contexts. In this paper, we describe the set-up of the CAUCP and the main features of the dataset and we present promising research prospects.

## 1. Introduction: Coronavirus crisis & public opinion

This article introduces data collected in the Citizens’ Attitudes Under Covid-19 Project (CAUCP), which surveyed public opinion throughout the Covid-19 pandemic in 22 countries^1^ from March to December 2020. In this paper, we present the dataset collected in 11 countries, which is deposited in open access on the Sciences Po Dataverse. It constitutes a unique cross-country panel survey of citizens’ attitudes and behaviors during a worldwide unprecedented health, governance, and economic crisis. Because this crisis tapped into all social dimensions, or *crisis characteristics*, it affected most peoples’ life (van der Ven and Sun 2021). The Covid-19 crisis has evoked seldom seen before individual and collective challenges to the public’s health, economy, and social life. The pandemic has potentially reshaped democratic politics in general and, in particular, affected attitudes on a large range of issues and influenced political behaviors. The investigation of citizen’s adaption and response to this crisis, as well as their (possibly enduring) consequences is interesting for several reasons. While the Covid-19 pandemic is no ordinary crisis affecting all dimensions of social life, it resonates with established findings on public opinion in times of crisis. First, according to the responsiveness and accountability perspectives (Soroka and Wlezien 2010) governments’ response to the crisis is conditioned by public opinion. Specifically, in the case of the Covid-19 pandemic, citizens’ (expected) preferences and partisanship have shaped governments’ response to the virus, sometimes at the expense of scientific expertise (Adolph et al. 2020). However, while the level of political trust has been linked to some policy responses, most studies evaluating them seem to overlook public opinion – mostly because of a lack of reliable and comparative data (Petridou 2020, Greer et al. 2020). Second, citizens’ reaction to Covid-19 related policies are also conditional on citizens’ political attitudes as well as emotions associated with the crisis. Particularly, sociodemographic characteristics, personality traits, ideology and emotions influence compliance to public health measures (Brouard et al. 2020). Third, the Covid-19 pandemic has shattered economies and produced a globalized economic crisis of major magnitude (OECD 2020). Economic crises, their perception, and the perception of governments’ responsibility normally have significant electoral and attitudinal consequences (Lewis-Beck and Stegmaier 2000, Hernandez and Kriesi 2016).

### Overview of existing knowledge/data on public opinion

The unprecedented crisis triggered by the Covid-19 pandemic has generated an enormous amount of survey research. However, because of limitations in resources, narrow methodologies, or absence of a comparative perspective, few studies allow an assessment of the transformations (or stability) of public opinion from the outbreak of the crisis in March 2020 onwards. For instance, available cross-country surveys are mostly cross-sectional, thus overlooking the evolutions of public opinion, and the durability of the effect, throughout a (at least) year-long eventful crisis (e.g. the ten-country study of Dryhurst et al. 2020). On the other hand, necessary panel studies are often limited to a single country, thereby overlooking the evolution of public opinion across a variety of contexts (see Kittel et al. 2020 for Austria and Hegewald and Schraff 2020 for the Netherlands, or the critique of limited geographical scope in Devine et al. 2020). Some studies do combine cross-country comparisons with panel designs through opt-in questions added in already existing panel surveys – although necessarily limiting the breadth of investigation (e.g. Ares et al. 2020). Addressing all these shortcomings, the CAUCP provides public opinion panel data from 11 democracies over the year 2020 in order to examine the longitudinal and cross-country evolution (or stability) of citizens’ preferences, perceptions and behaviour.

## 2. Citizens’ Attitudes Under the COVID-19 Pandemic: generating a unique dataset

### 2.1. Research questions

The CAUCP cross-country panel survey sets to answer four broad research questions, covering four major dimensions of the effects of the Covid19 pandemic on public opinion. First, **[RQ1] Electoral Behavior** examines the electoral consequences of the Covid-19 pandemic to evaluate its effects (if any) on prospective electoral behavior (turnout, vote choices and vote intentions), as well as the influence of retrospective vote choice on behaviors and attitudes linked to the Covid-19 crisis. Second, **[RQ2] Political Behavior and Attitudes** investigates how people respond to the crisis in terms of political preferences, political trust and satisfaction, policy evaluation, blame attribution, emotions, and more generally preferences on the role of the State. Third, **[RQ3] Behavioral and attitudinal reaction to Covid19 related policies** sets to explain the reasons why people are more or less likely to support and comply with public Covid-19 related recommendations and under which circumstances. The survey also examines how people acquire information about the Covid19 pandemic. It addresses the causal mechanisms by which people perceive the salience of the pandemic and how they react to the discourse of public authorities, media outlets, and other public opinion leaders. Fourth, **[RQ4] Social Consequences** examines the conditions under which the COVID-19 crisis increases fragmentation within societies and affects social cohesion. The survey examines the dynamics of individual economic well-being, social isolation, occupational trajectories, and patterns of socialization.

## 3. Study design and dataset description

### 3.1. Study design

Based on 11 democracies, the rationale of case-selection combines a variety of characteristics: geographical scope, intensity of the pandemic and timing of its development, type and stringency of the policy response, institutional characteristics (member of the EU or not, centralized vs. federal governance). The CAUCP panel data was collected (Computer-Aided Web Interview) in four waves, from the outset of the pandemic (Wave 1 in late March 2020) and throughout 2020 (Wave 2 in April 2020, Wave 3 in June 2020, and Wave 4 in December 2020). Therefore, the first wave of the panel fits closely with the implementation of the first health restrictions in mid-March in Europe. The data collection covers the evolution of the pandemic over the summer (lower intensity of the pandemic and loosened restrictions in the Northern hemisphere vs. higher intensity of pandemic and more stringent policies in the Southern hemisphere), and it extends to December 2020 (as the pandemic dynamic reverses again between South and North). Figure 1 summarizes the fieldwork information (countries included, date of data collection, number of respondents)^2^.

**Figure 1 -.**
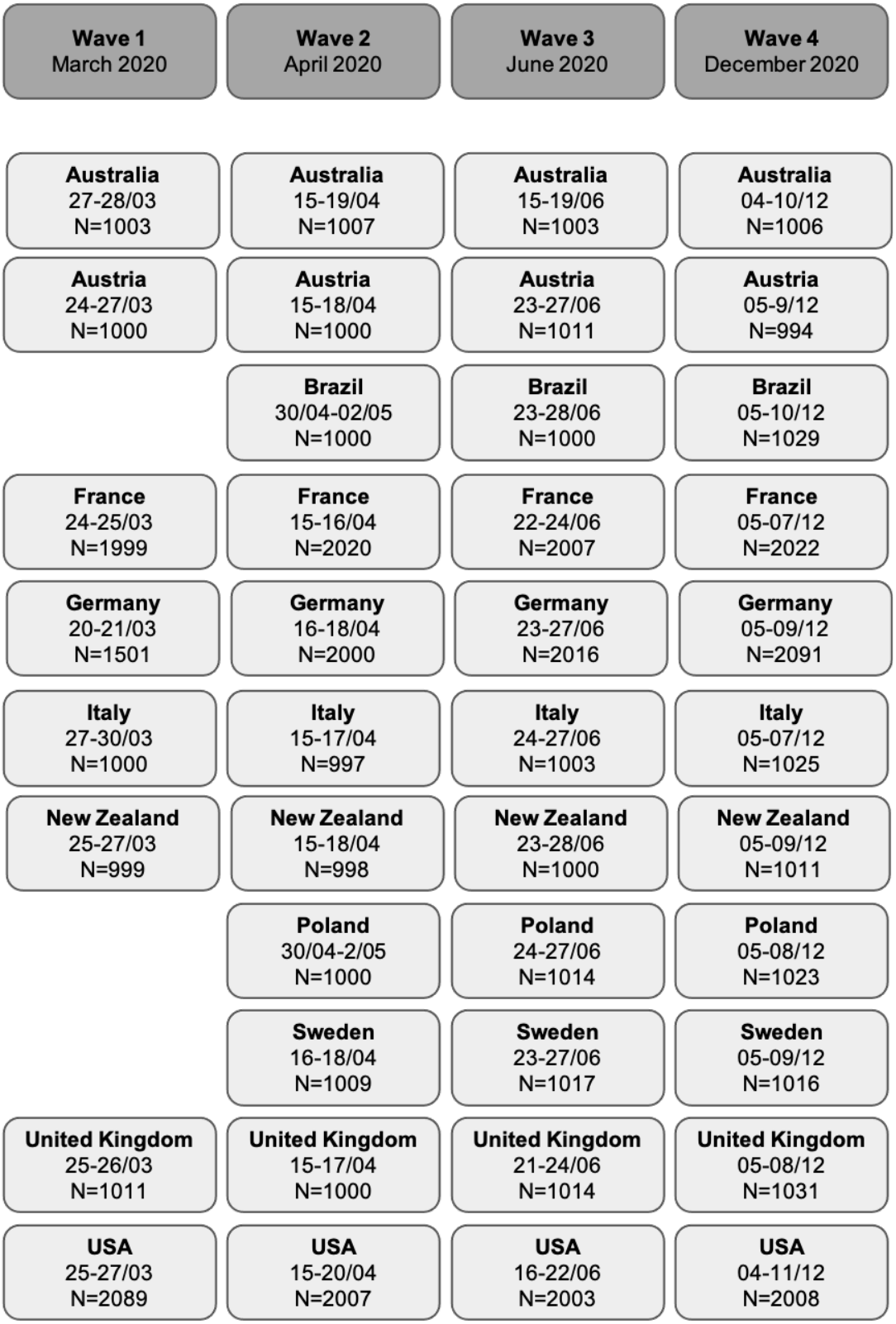
Design of the cross-country CAUCP panel study.

### 3.2. Survey content: topic overview of variables

Due to space restrictions, in the current article we cannot present the exhaustive list of variables from the CAUCP surveys as it exceeds 300 variables across countries and waves. However, Table 1 presents a broad overview of most relevant topics and variables corresponding to the four aforementioned research questions.^3^

**Table 1 -.**
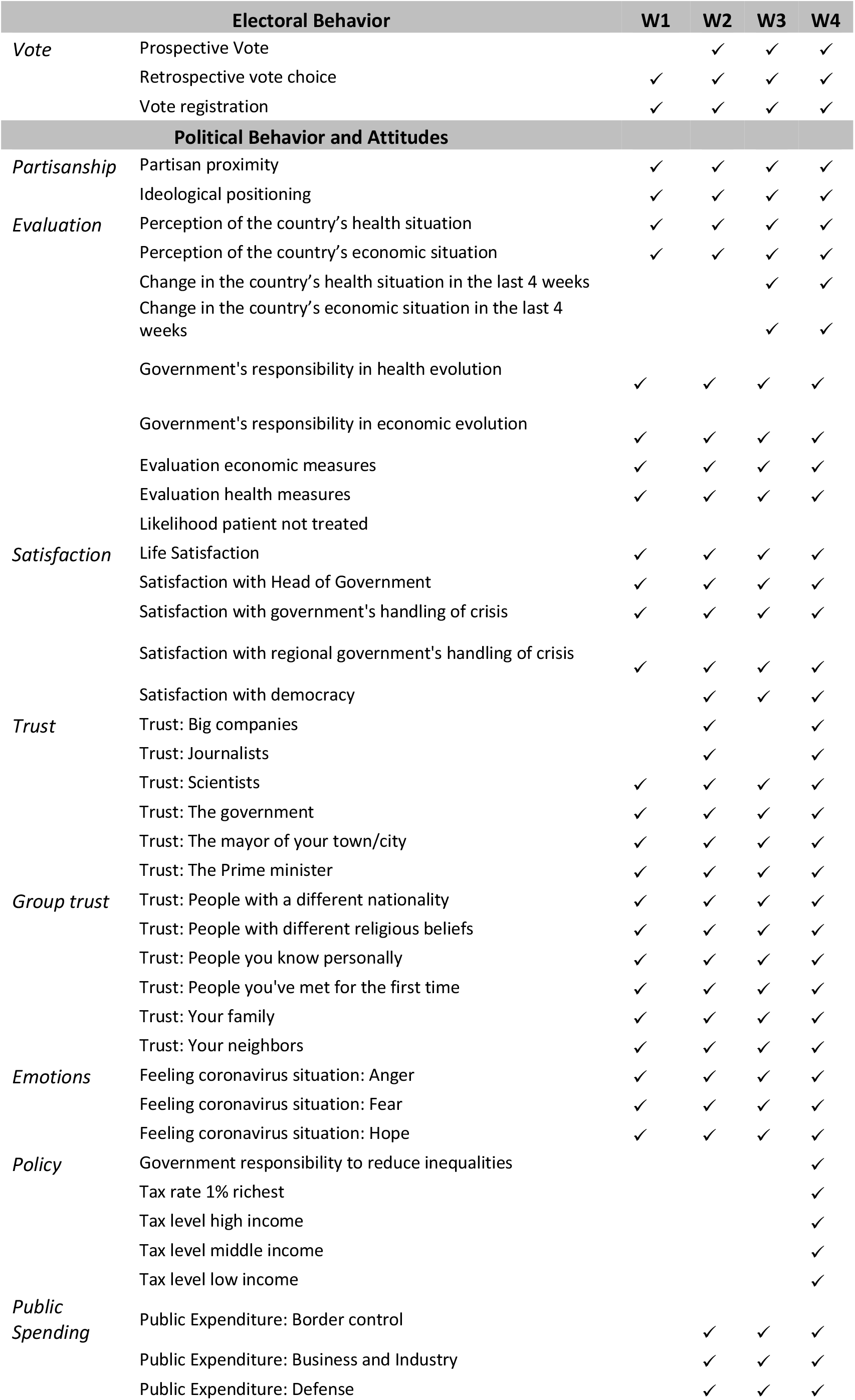
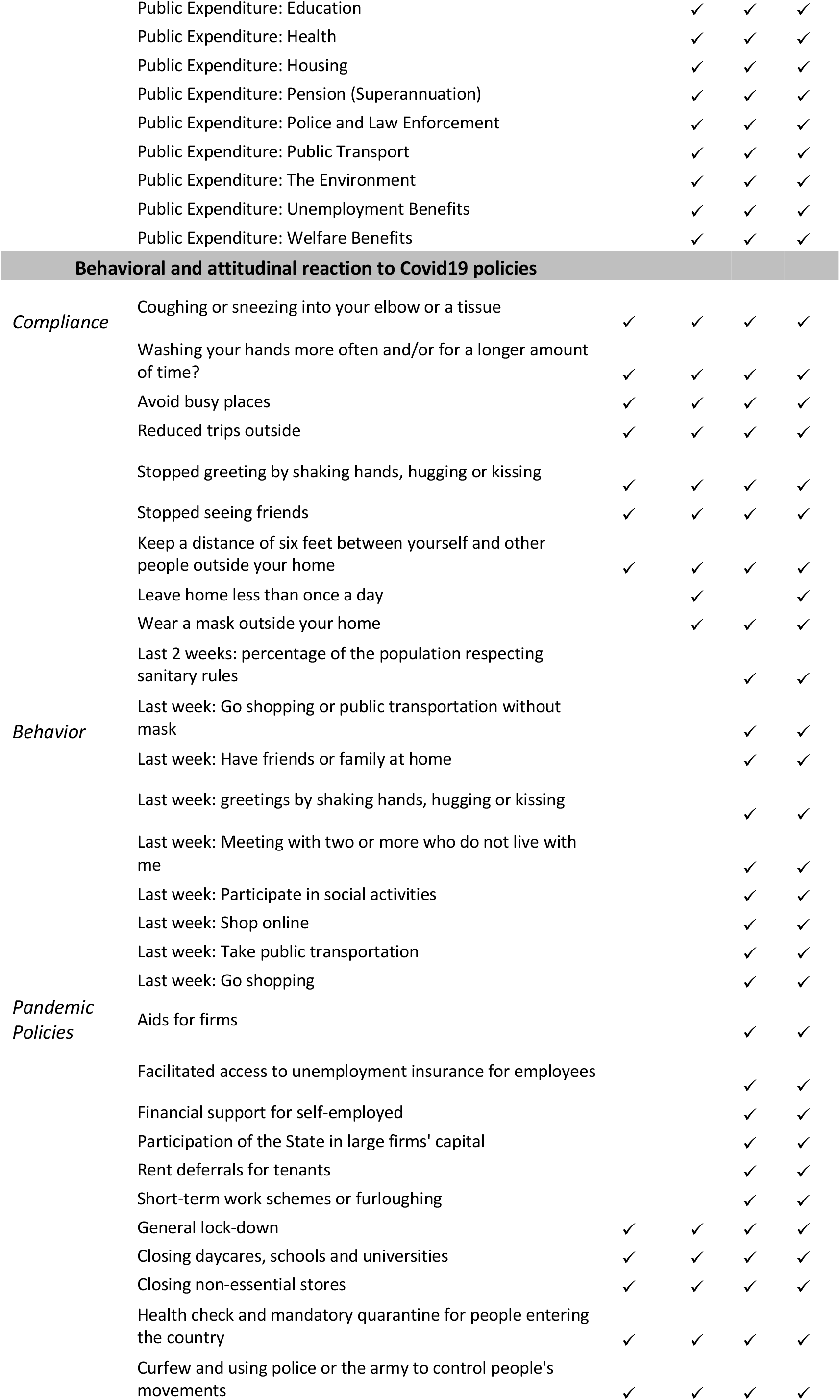
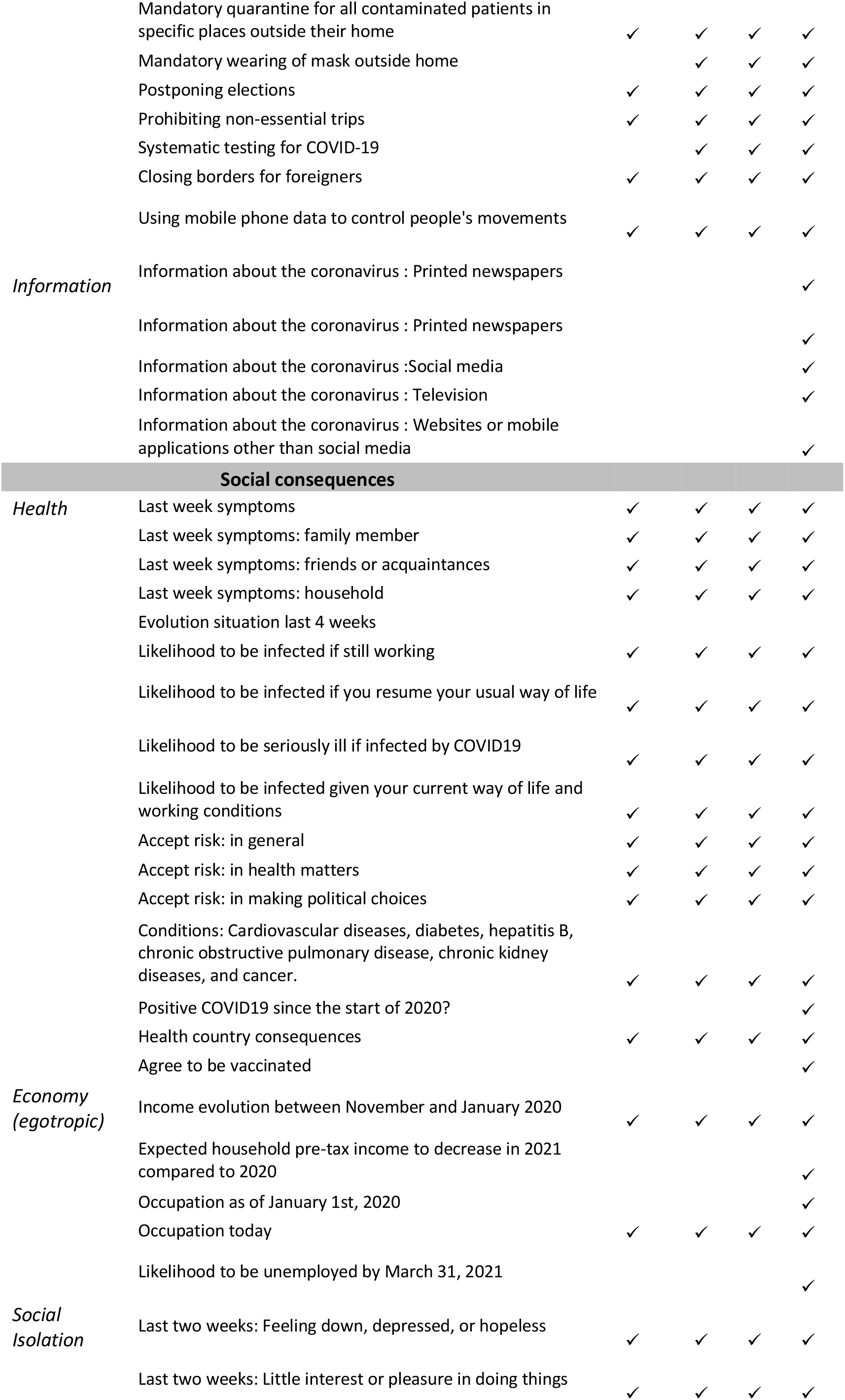
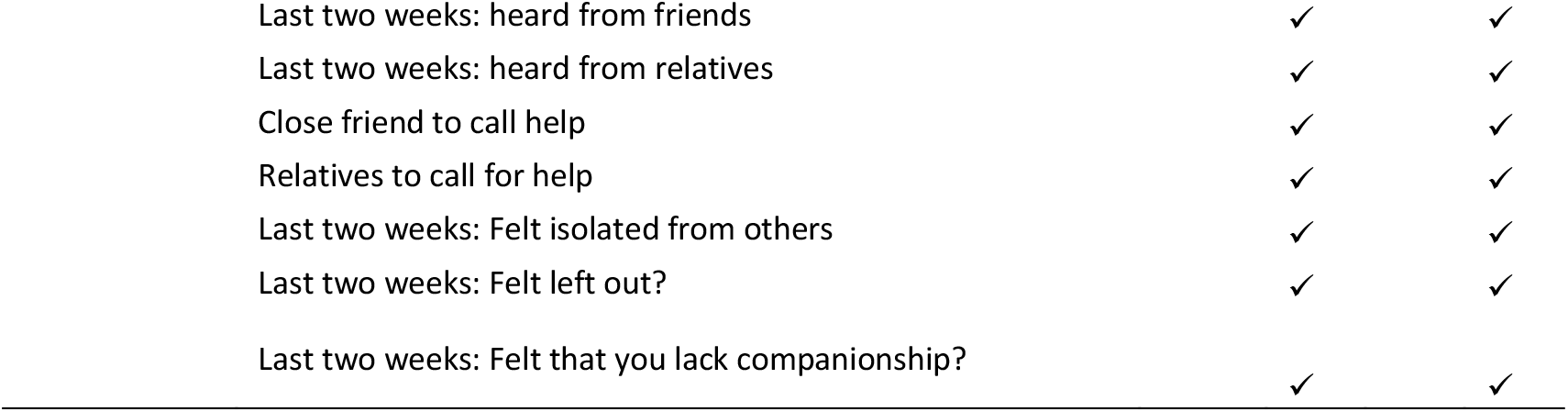
Scope and main variables of interest of the CAUCP survey.

#### 3.3. Data quality

A minimum of 1,000 respondents were surveyed in each panel wave (and about 2,000 in France, Germany, and the USA) ensuring representative samples with quotas. The questionnaire retained most questions throughout the waves, to allow the longitudinal analysis of variables. However, questionnaires were flexible between waves and countries in order to include newly salient topics (e.g. “wearing masks” from wave 2) and to adapt to the countries’ specific contexts (e.g. questions on multi-level governance, or national/local elections). The CAUCP included a total of 25,569 panelists residing in the country where they are surveyed. Despite the period of 10 months between Wave 1 and Wave 4 (the share of respondents who completed the four waves ranges from 20% to 50% - Figure 2).

**Figure 2 -.**
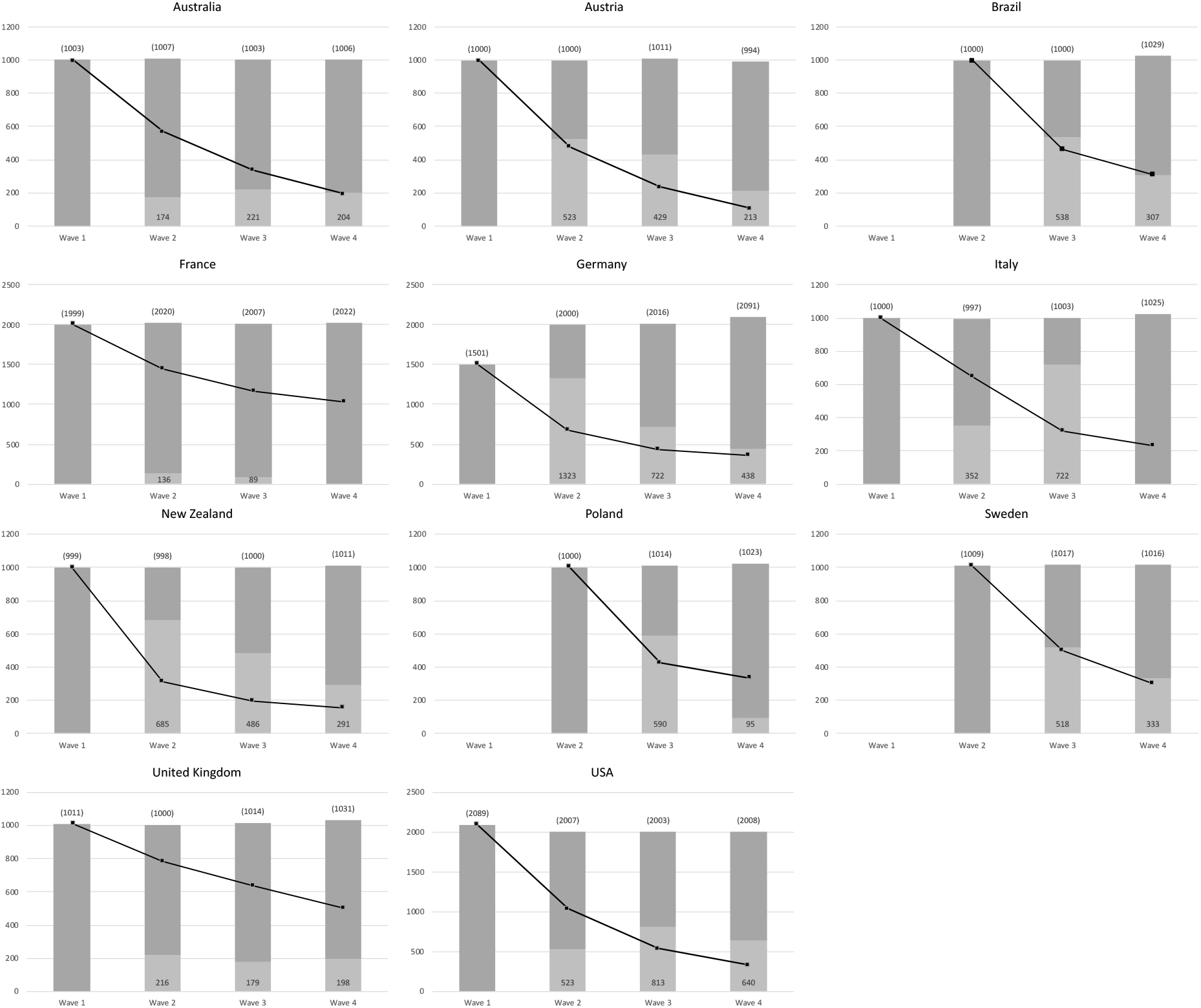
Number of respondents and attrition of panelists. Light grey areas indicate new respondents in a Wave. Black line reports the number of respondents included in all four waves.

## 4. Research prospects

In this section, we highlight how the CAUCP data may be useful to enrich a variety of social research areas. Particularly, we present three key dimensions covered by our survey: “democratic accountability”, “policy compliance” and “attitudinal dynamics”. We highlight some key variables to study the possibly large and lasting effects of the Covid-19 crisis on social and political life.

### 4.1. Democratic accountability

Studying the consequences of the Covid19 pandemic in terms of democratic accountability represents a broad research agenda. Citizens’ response to the unprecedented pandemic policies, and to the subsequent expected economic crisis, is crucial to understand the functioning of democracies and will most likely be multifaceted (electoral behavior, trust and evaluation of institutions and policies, blame attribution). For instance, initial findings on citizens’ response to the Covid-19 pandemic show a ‘rally around the flag effect’ (Baekgaard et al. 2020). However, this effect should be qualified since it is neither universal nor similarly lasting as the crisis endures (Herrera et al. 2020), or rather producing a ‘*crisis signal effect that benefits incumbents*’ (in Italy, De Vries et al. 2020). While earlier findings suggest that the pandemic has triggered stronger support or satisfaction with the government, they mostly rely on single-country studies. The CAUCP data suggest that such policy response is more complex and context dependent. Looking at the satisfaction with the government’s handling of the Covid19 crisis, we observe important time and country variation (Figure 3). Indeed, in the different countries, the level of satisfaction is extremely high (Australia, New Zealand, Austria), average (France, Italy, USA), or generally low (Brazil, Poland). Additionally, trends of satisfaction are either increasing (Australia), stable (France, New Zealand), or declining (Sweden, United Kingdom). Beyond the way by which citizens evaluate their government, it raises some questions on the electoral consequences of the crisis: are these patterns consistent? Are they dependent on the context of the pandemic and the policy response? Do they hold for national and local elections? Such questions enable to investigate promising research venues on the effects of retrospective vote choice on attitudes, as well as on the consequences of the crisis on prospective vote choice, and actual electoral behavior as three countries held national elections in 2020 (USA, New Zealand, and Poland). Evaluating the significance of individual-level health and economic situations on political behavior and attitudes constitute a promising research lead, as most studies examined these questions at the aggregated level thus far.

**Figure 3 -.**
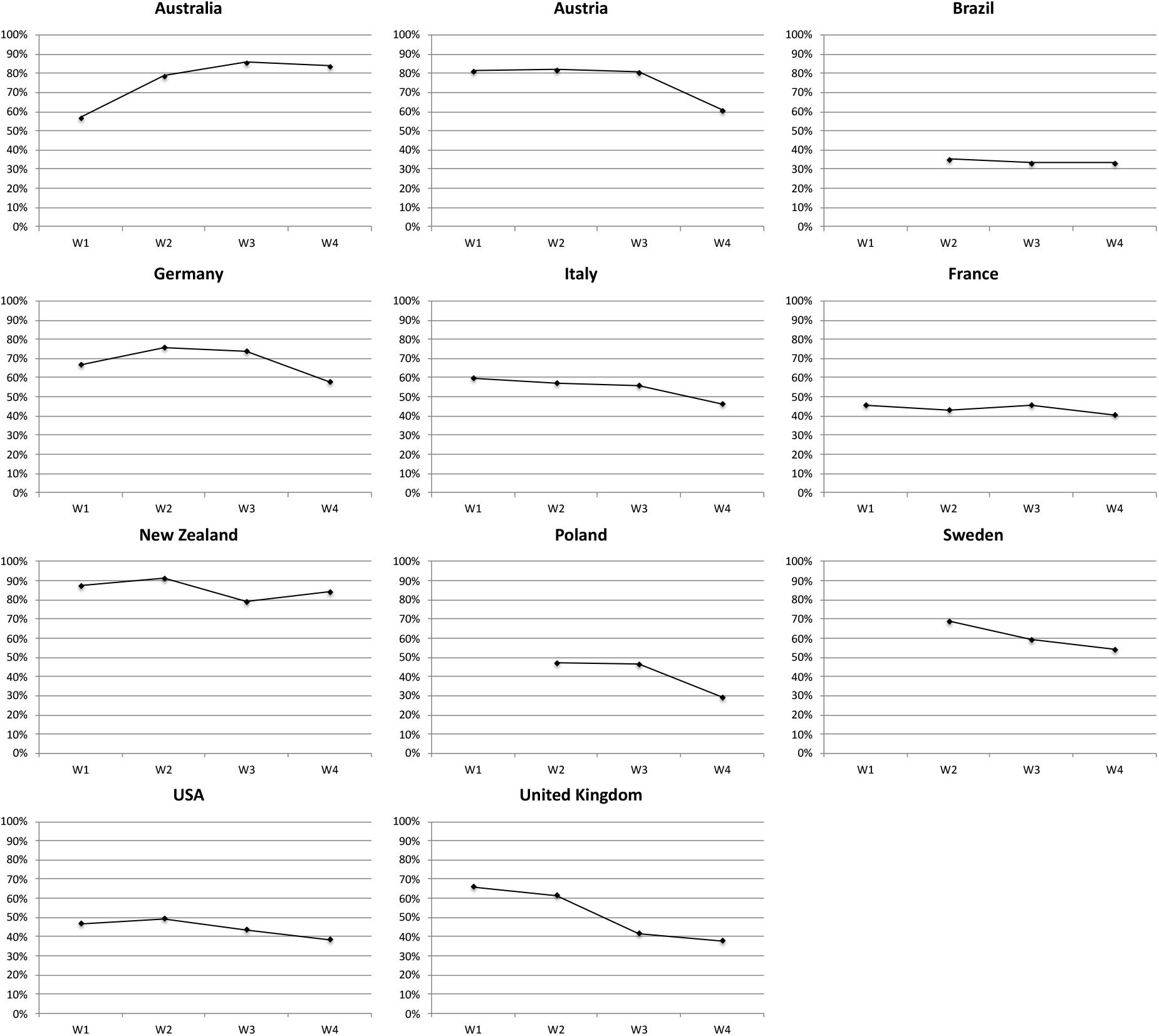
Satisfaction with the government’s handling of the Covid19 crisis^*4*.^

### 4.2. Politics of policy compliance

Compliance with Covid-19 related policies is both a salient political debate and constitutes a promising research agenda. Indeed, the preventive and sometimes restrictive policies implemented during the Covid-19 pandemic have been debated and to a large extent approved and respected by citizens. We identify two dimensions of *policy compliance*: individual behavior (i.e. respecting restrictive regulations) and acceptance/evaluation of Covid-19 related measures (i.e. approval of policies limiting rights and liberties). Despite sharp political volatility, such as decline (or variations) in trust and negative evaluations of the government, compliance levels to stringent policies remained high (Newton 2020). However, compliance is mediated by partisanship: support for the parties in government leads to greater compliance (for the USA Barrios and Hochberg 2020, for Belgium Dyevre and Yu-Cherong Yeung 2020). The politics of complying to stringent health policies is certainly influenced by individual and contextual factors. For instance, patterns of compliance to ‘keeping social distance’ diverge across countries, but also evolve over time (Figure 4). Even in the case of New Zealand which has been less exposed to the coronavirus in terms of fatalities, citizens do react to the Covid-19 related policies and adapt to government recommendations. Indeed, pandemic policies are the most visible policies: virtually all citizens are aware and informed on Covid-19 restrictions. This constitutes a unique research area to evaluate both individual and contextual determinants of policy acceptance.

**Figure 4 -.**
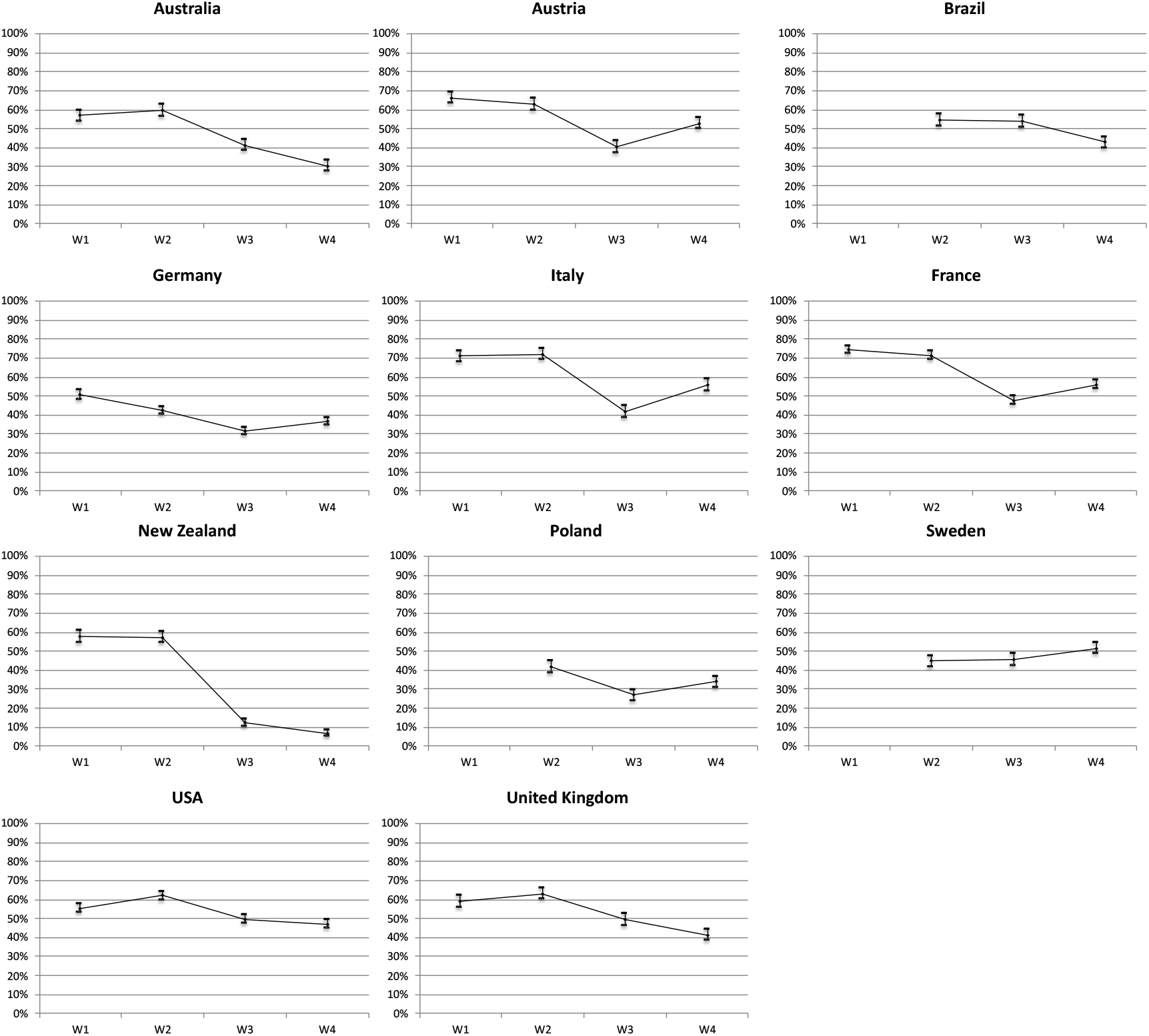
Compliance: Keeping distance with people outside home^5^.

### 4.3. Attitudinal dynamics

The Covid-19 pandemic has had consequences for public attitudes. Some speculate that the Covid-19 may mark a ‘great transformation’ that will deeply modify political cleavages. The pandemic may also polarize or de-polarize democratic societies. Alternatively, the political impact of the pandemic may be sectoral only, that is, limited to the specific dimensions linked to the crisis. Finally, the Covid-19 pandemic might not be a more influential crisis, as long as citizens’ attitudes are concerned, compared to previous major crises (geopolitical shocks, economic crisis). Still, findings suggest that the pandemic has triggered important attitudinal changes, which however mask individual-level heterogeneity within each country of our sample (for instance Galasso et al. find sizeable gender differences in attitude during the crisis). What are the factors driving these attitudinal changes? As new issues appeared on the public agendas, there are also opportunities to study the formation of attitudes. Furthermore, many psychological traits, such as risk perception, emotions or socialization are expected to greatly influence political attitudes. Even more, the enormous amount of (mis-) information on the Covid-19 pandemic most likely influenced psycho-political attitudes. Indeed, earlier findings have found evidence that different patterns of news consumption affected levels of trust and compliance to Covid-19 related restrictions (Newton 2020). Yet, in terms of specific economic beliefs, early data suggests that the economic crisis resulting from the pandemic seems to have almost no impact (Ares et al. 2020). Descriptive findings of the CAUCP survey tend to indicate that the pandemic has not transformed overall political preferences. Indeed, average ideological preferences (Left-Right 11-point scale) are stable over the waves of the panel, and polarization seems unchanged (roughly measured with the standard deviation of the Left-Right scale). Despite a political anchor such as ideology remaining stable throughout the crisis other attitudes tend to fluctuate across time and contexts. For instance, preferences for closing borders have changed during the crisis, although in contrasted directions (Figure 5). Even further, the political consequences of the pandemic seem to be mediated by anxiety rather than a cognitive trigger of the political ‘rally’ effect (Scharff 2020, Kritzinger & al. 2021). The CAUCP offers a promising agenda for research in political psychology with a large battery of questions on emotions and social interactions.

**Figure 5 -.**
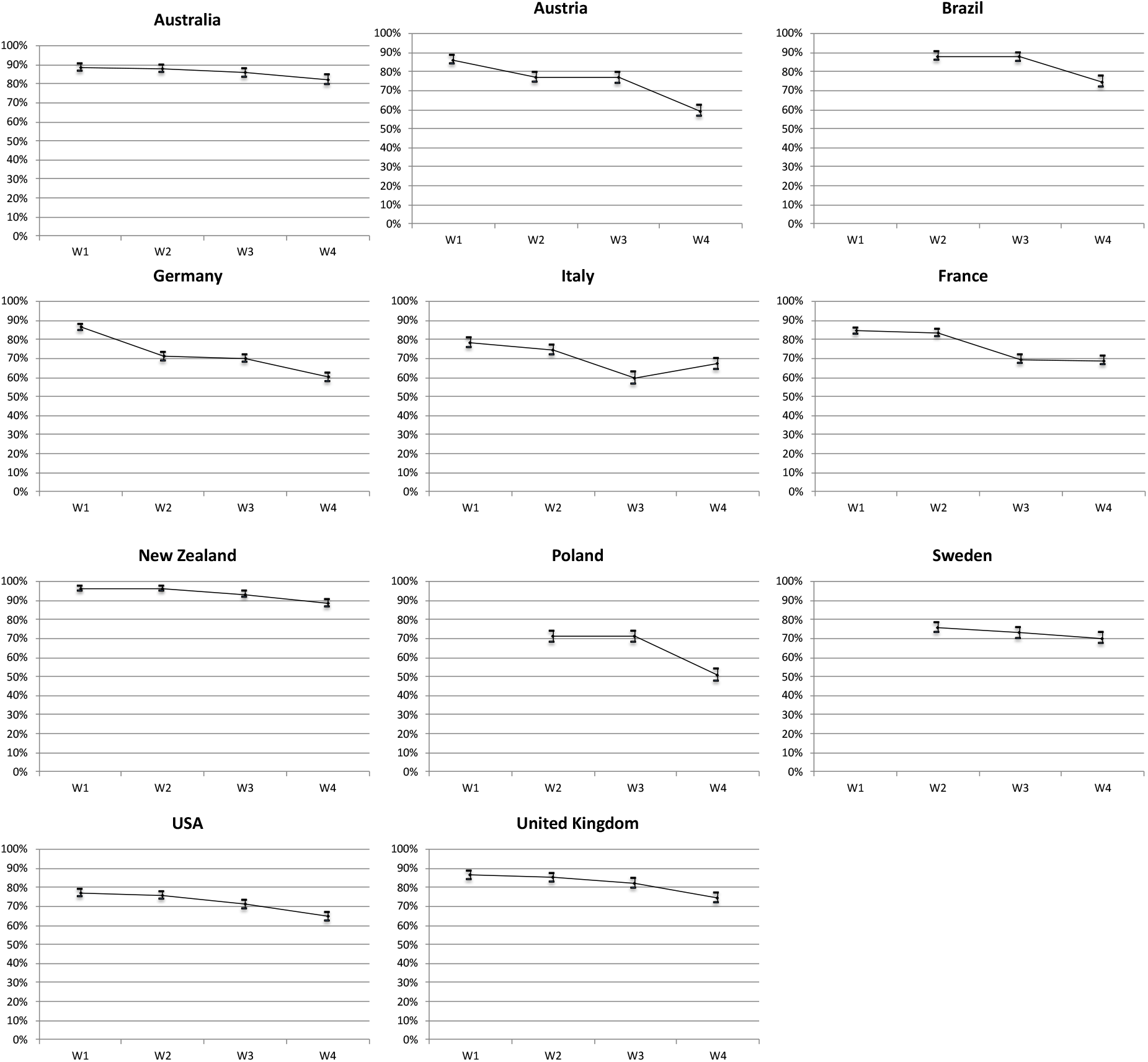
Preferences for closing the borders to foreigners^6^.

## 5. Data accessibility and conclusion

The CAUCP data provides unique information on public opinion throughout the Covid-19 pandemic. It allows to examine the behavioral and attitudinal consequences of this major health, economic, and social crisis across time and contexts. In this paper, we have described - 5-point Likert Scale. Figure 5 shows the proportion of respondents who “completely agree” or “tend to agree”. the set-up of the CAUCP, and the main features of the dataset. We only hinted at some aspect of the vast scientific agenda that this data can inform. In order to encourage further research on public opinion in the time of the Covid19 pandemic, the CAUCP dataset is deposited on the Sciences Po Dataverse and accessible in open access using the below link. We have uploaded sub-datasets for each wave in each country of the study in TAB format. The country*wave sub-datasets have been cleaned to standardized variable names across country and wave subdatasets datasets. We have also included detailed codebooks for each for each sub-dataset which describes all the variables included in each sub-dataset (variables labels, answer categories, answer labels, country specific variables) in XLSX format. All this material is available at the following link: https://data.sciencespo.fr/dataset.xhtml?persistentId=doi%3A10.21410%2F7E4%2FEATFBW

In addition to the individual country*wave files and documentations uploaded on the institutional repository of Sciences Po, we have uploaded a merged dataset on figshare repository. This merged dataset includes the observations for the eleven countries and the 4 waves, as well as an extended codebook with (variables types, names, labels, values, and value labels) in XLSX format.

## Data Availability

We have uploaded sub-datasets for each wave in each country of the study in TAB format. The country*wave sub-datasets have been cleaned to standardized variable names across country and wave subdatasets datasets. We have also included detailed codebooks for each for each sub-dataset which describes all the variables included in each sub-dataset (variables labels, answer categories, answer labels, country specific variables) in XLSX format. All this material is available at the following link:
In addition to the individual country*wave files and documentations uploaded on the institutional repository of Sciences Po, we have uploaded a merged dataset on figshare repository. This merged dataset includes the observations for the eleven countries and the 4 waves, as well as an extended codebook with (variables types, names, labels, values, and value labels) in XLSX format.

https://data.sciencespo.fr/dataset.xhtml?persistentId=doi%3A10.21410%2F7E4%2FEATFBW

## Footnotes

1 Argentina, Australia, Austria, Burkina-Faso, Brazil, Canada, Egypt, France, Germany, Italy, Ivory Coast, Maroc, New Zealand, Niger, Nigeria, Poland, Senegal, Sweden, South Africa, Spain, United Kingdom, United States of America.

2 Because of technical and resource constraints, three countries were included in the data collection starting from Wave 2 only (Brazil, Poland, and Sweden).

3 In addition to the variables presented in Table 1, the CAUCP survey includes extensive information on respondents’ socio-demographic characteristics (gender, age, religiosity, income, education, household description, employment status and trajectory). In this overview, the variables related to a series of randomized experiments are also excluded. Particularly, such experiments focused addressed RQ3 and focused on individual compliance to health regulations – data and results are presented separately elsewhere.

4 Question wording: “Generally speaking, are you satisfied with the way that the government is handling coronavirus?” – 4-point Likert Scale. Figure 3 shows the proportion of respondents that are “completely satisfied” or “quite satisfied”.

5 Question wording: **“***Due to the coronavirus epidemic, in your daily life, would you say that you keep a distance of three feet (1 meter) between yourself and other people outside your home? 0 means “not at all” and 10 means “yes completely***”. Figure 4 shows the proportion of respondents who score 10. We consider that the latter respondents are those expressing full compliance to restrictive measures**.

6 Question wording: “*Here is a list of measures that have been taken in some countries against the spread of coronavirus (N-Covid19). Do you agree with them? The closing of the borders for foreigners*”

